# COVID-19 Clinical Characteristics and Outcomes in Children and Adolescents Hospitalized at the University Hospital of the West Indies, Jamaica in 2020-2021

**DOI:** 10.1101/2021.11.26.21266916

**Authors:** Crista-Lee Shahine Berry, Roxanne Helene Melbourne-Chambers, Abigail Natalie Harrison, Joshua James Anzinger, Kelly-Ann Maxorinthia Gordon-Johnson, Varough Mohamed Deyde, Celia Dana Claire Christie

## Abstract

**Background and Objectives:** Multisystem inflammatory syndrome of children (MISC) carries a high attributable morbidity. We describe children aged <16 years hospitalised with COVID-19 and/or MISC, April 2020 to June 2021.

**Methods:** All were tested for SARS-CoV-2, infectious disease consultations performed, modified CDC criteria for MISC applied, charts reviewed and data analyzed.

**Results:** Among 79 consecutive children with SARS-CoV-2, 41(52%) were hospitalised; with median age 10.5 years; Afro-Caribbean ethnicity 40(98%); males 21(51%); SARS-CoV-2 RT-PCR positivity 26 (63%), IgG/IgM positivity 7(17%), community exposures 8 (20%). MISC-cases 18 (44%) vs. non-MISC 23(56%) had fever (94% vs. 30%; p<0.01), fatigue/lethargy (41% vs. 4%; p=0.004), rhinorrhoea (28% vs. 4%; p=0.035), elevated neutrophils (100% vs. 87%; p=0.024) and ≥4 abnormal inflammatory biomarkers 13 (72%). MISC-cases had ≥2 organ/systems (100% vs. 35%; p<0.01), including gastrointestinal (72% vs. 17%; p<0.01), haematological/coagulopathic (67% vs. 4%; p<0.01); dermatologic (56% vs. 0%; p<0.01), cardiac (17% vs. 0%; p=0.042) with Kawasaki Syndrome (44% vs. 0%; p<0.01) and pleural effusions (17% vs. 0%; p=0.042). MISC-cases were treated with intravenous immune gammaglobulin (14, 78%), aspirin (12, 68%), steroids (9, 50%) and intensive care with non-invasive ventilation (2, 11%). One (6%) with pre-morbid illness died, the remainder recovered.

**Conclusion:** MISC was treated successfully with intravenous gammaglobulin, steroids and/or aspirin in 94% before cardiopulmonary decompensation, or need for inotropes, vasopressors, or invasive ventilation.

## BACKGROUND

Since the detection of SARS-CoV-2, the virus causing COVID-19 in Wuhan, China in December 2019, it rapidly spread with a pandemic declared in March 2020 (World Health Organization Director General, 2020). Initially, children were asymptomatic, or had a milder course with better prognosis than adults (Joseph et al., 2020). By April 2020, the post-infectious Multisystem Inflammatory Syndrome in Children with COVID-19 (MISC), the Paediatric Multisystem Inflammatory Syndrome (PIMS), began to manifest in children in several countries (Miller J et al., 2020; Royal College of Paediatrics and Child Health, 2020; Centres for Disease Control and Prevention, 2020; World Health Organization, 2020).

Three diagnostic criteria for MISC/PIMS have been created by the Centers for Disease Control and Prevention (CDC), World Health Organization (WHO) and the Royal College of Paediatrics and Child Health, (RCPCH). The CDC’s criteria includes patients <21 years with objective or subjective fever for ≥24 hrs, laboratory evidence of inflammation (≥one: elevated C-reactive protein, erythrocyte sedimentation rate (ESR), fibrinogen, procalcitonin, d-dimer, ferritin, lactic acid dehydrogenase, interleukin-6, elevated neutrophils, reduced lymphocytes and low albumin) and clinically severe illness requiring hospitalization, with multisystem (≥2) organ involvement (cardiac, renal, respiratory, hematologic, gastrointestinal, dermatologic or neurological); no alternative diagnoses; positive for current, or recent SARS-CoV-2 infection by RT-PCR, serology, or antigen test; or exposure to a suspected, or confirmed COVID-19 case in 4 weeks prior (Centers for Disease Control and Prevention, 2020). Children meeting full or partial criteria for Kawasaki disease, or any paediatric death with SARS-CoV-2 infection is also considered MISC.

The WHO’s criteria include patients aged <19 years with fever for three days, two organ systems involved, elevated inflammatory markers; no other microbial cause for inflammation and SARS-CoV-2 infection, or exposure (World Health Organization, 2020). The RCPCH has persistent fever, inflammation, organ dysfunction, an alternative microbial cause excluded and SARS-CoV-2 RT-PCR positivity, or negativity (RCPCH, 2020). The American Academy of Paediatrics recommended a treatment approach for children with MISC (American Academy of Paediatrics, 2021).

Jamaica, a middle-developing, Caribbean Island nation of 2.9 million inhabitants, diagnosed its first imported COVID-19 case on March 10, 2020; a day before COVID-19 was declared a pandemic (COVID-19 Ministry of Health, Jamaica, 2020; Christie CDC et al., 2020). Face-to-face school attendance was discontinued and virtual classes began approximately 5 weeks later. Strict public health measures, including mandatory mask wearing in public spaces, physical distancing, sanitizing, closure of the island’s borders and varied “stay-at-home” orders were implemented. On June 1, 2020, Jamaica eased lockdown restrictions and on June 15, 2020, international borders were reopened to airline passengers. Community transmission was declared in September 2020 with positivity rates fluctuating between 8% and 41%, peaking in March 2021. Concerns about learning loss in children, stimulated discussions on the timing and safe reopening of schools. Hence, description of the clinical characteristics and severity of outcomes of the COVID-19 in Jamaican children is of utmost importance.

As of June 1^st^ 2021, Jamaica reported 48,639 COVID-19 confirmed cases; 10.3% (4999) were aged <18 years. Five percent (250) were under one year old, 10.4% (520) were aged 1-2 years, 13.1% (653) 3-5 years, 36.1% (1807) 6 – 12 years and 35.4% (1769) 13 -17 years old. Deaths were reported in 0.1% under 10 years and 0.2% 10-19 years old (9). The Caribbean Public Health Agency reported the B.1.1.7 (alpha, UK) SARS-CoV-2 as the only “variant of concern” identified in Jamaica through the third week of June, 2021 (Charles J, 2021; COVID-19 Ministry of Health, Jamaica, 2021). The co-circulation of the B1.617.2 (Delta, Indian) “variant of concern” has been reported since June 23, 2021 (COVID-19 Ministry of Health, Jamaica, 2021; PAHO/WHO, 2021).

The University Hospital of the West Indies (UHWI), Jamaica, is a tertiary academic institution that accepts referred and ambulatory adult and paediatric patients island-wide and from the English-speaking Caribbean, including for intensive care services. SARS-CoV-2 RT-PCR nasopharyngeal testing is done on every admitted patient and SARS-CoV-2 antibody testing, when indicated. Although SARS-CoV-2 RT-PCR has very high analytical sensitivity, diagnostic sensitivity is imperfect and false negatives range from 1.8%-58% (Arevalo-Rodriguez et al., 2020). Use of SARS-CoV-2 RT-PCR followed by antibody testing increases diagnostic sensitivity for disease occurring after clearance of virus from the upper respiratory tract (Wang et al., 2020).

We describe the clinical characteristics, therapy and outcomes in patients aged <16 years old admitted to UHWI with SARS-CoV-2 infection, or MISC. This enabled improved clinical identification and development of best practices for SARS-CoV-2 infection in children and contributes to public policy in Jamaica and global health knowledge of children with SARS-CoV-2 infection and MISC for developing countries.

## METHODS

This descriptive, ambispective study included all patients aged <16 years, with SARS-CoV-2 infection presenting to the UHWI and those meeting the CDC’s diagnostic criteria for MISC and non-MISC cases, hospitalized at the UHWI, Jamaica during April 1, 2020 through June 30, 2021. Utilising careful chart review, we describe clinical characteristics, investigations, treatment and outcomes of the population.

All hospitalized patients have mandatory nasopharyngeal RT-PCR testing for SARS-CoV-2. All suspected and confirmed patients had a Paediatric Infectious Diseases consult and upon discharge, were reviewed by the paediatric inpatient, or outpatient department. SARS-CoV-2 infection was confirmed by RT-PCR positivity of nasopharyngeal samples at the National Influenza Centre, presence of IgG and/or IgM antibodies (Abbott Architect), and/or confirmed epidemiological link, or presumed contact with an infected person. Electronic and physical clinical records were reviewed by senior researchers and a clinical research assistant. Clinical features, investigations, treatments and outcomes were extracted using a modified WHO case report form and data were entered into RedCap database by senior clinical researchers.

Statistical analyses were performed using IBM SPSS version 28. Descriptive statistics were obtained for clinical characteristics and outcomes in children <16 years old on admission using mean and standard deviation for continuous variables and number and percent for categorical variables. Comparison of differences between children with, or without MISC was computed using t-tests for continuous variables and chi-squared test for categorical variables. A p value <0.05 was considered statistically significant.

## RESULTS

During April 2020 to June 2021, 79 children and adolescents, aged <16 years, presented to the UHWI with SARS-CoV-2 RT-PCR positive status, or met the CDC’s criteria for MISC. Of these, 41(52%) children were hospitalized (median age 10.5 years; range 0.003–15.92 years); SARS-CoV-2 RT-PCR positivity present in 26(63%); SARS-CoV-2 IgM/IgG positivity in 7(17%), confirmed/presumed community contact in 8(19.5%) (Table 1). Twenty-one (51.2%) were male and 40(98%) were of Afro-Caribbean ethnicity. Twenty-two (53.7%) had co-morbidities with sickle cell disease (6; 27.3%), immunodeficiency (4;18.2%) and asthma (6; 27.3%) being of greatest prevalence.(Table 1)

**Table 1:** Characteristics of children hospitalized with COVID-19, MISC* vs non-MISC.

|  | **Total cases**<br>**(%) N= 41** | **MISC cases**<br>**(%) N=18** | **Non-MISC cases**<br>**(%) N=23** | **p-Value** |
| --- | --- | --- | --- | --- |
| **Male** | 21 (51.2%) | 9 (50%) | 12 (52%) | 0.890 |
| **SARS-CoV-2 RT PCR positive\*\*** | 26 (63.4%) | 4 (22.2%) | 22 (95.7%) | <0.001 |
| **SARS-CoV-2 IgG positive\*\*** | 6 (14.6%) | 6 (33.3%) | 0 (0%) |  |
| **SARSCo-V 2 IgM positive\*\*** | 4 (9.8%) | 4 (22.2%) | 0 (0%) |  |
| **Confirmed SARS-CoV-2 Contact** | 8 (19.5%) | 5 (27.8%) | 3 (13.0%) |  |
| **Suspected SARS-CoV-2 Contact** | 33 (80.5%) | 13 (72.2%) | 20 (87%) |  |
| **Infant (< 1 year)** | 8 (19.5%) | 1 (5.6%) | 7 (30.4%) | 0.041 |
| **Toddler (1 – 2 years)** | 6 (14.6%) | 4 (22.2%) | 2 (8.7%) |  |
| **Pre-schooler (3 – 5 years)** | 2 (4.9%) | 2 (11.1%) | 0 (0%) |  |
| Middle Childhood (6-11 years) | 9 (22%) | 6 (33.3%) | 3 (13.0%) |  |
| Adolescent (12-16 years)*** | 16 (39%) | 5 (27.8%) | 11 (47.8%) |  |
| **Comorbidities** | 22 (53.7%) | 9 (50%) | 13 (56.5%) | 0.577 |
| Congenital Heart Disease | 1 (4.5%) | 0 (0%) | 1 (7.7%) | 0.394 |
| Hypertension | 4 (18.2%) | 2 (22.2%) | 2 (15.4%) | 0.683 |
| Asthma | 6 (27.3%) | 3 (33.3%) | 3 (23.1%) | 0.595 |
| Chronic Kidney Disease | 2 (9.1%) | 1 (11.1%) | 1 (7.7%) | 0.784 |
| Chronic Lung Disease (not asthma) | 1 (4.5%) | 0 (0%) | 1 (7.7%) | 0.394 |
| Obesity | 3 (13.6%) | 2 (22.2%) | 1 (7.7%) | 0.329 |
| Epilepsy | 1 (4.5%) | 0 (0%) | 1 (7.7%) | 0.394 |
| Immunosuppression, Malignancy | 4 (18.2%) | 1 (11.1%) | 3 (23.1%) | 0.474 |
| Sickle Cell Disease, SCD | 6 (27.3%) | 4 (44.4%) | 2 (15.4%) | 0.132 |
| Other Comorbidities (Autism Spectrum D/O, Dysmorphic Syndrome, Hirschsprung's Disease, Metabolic Bone Disease, Sickle Nephropathy, Steroid Induced DM) | 8 (36.4%) | 3 (33.3%) | 5 (38.5%) | 0.806 |

\*Multisystem Inflammatory Syndrome in Children (MISC) with COVID-19, MISC.
\*\* RT-PCR was performed on ALL hospitalized patients. Antibody testing was not performed if the RT-PCR test was positive. Hence, no patient in the non-MISC group had antibody tests performed.
\*\*\*Children aged < 16 years are all admitted to the paediatric floors, those aged 16 years and older on presentation, are hospitalised on the adult floors. However one child celebrated his 16<sup>th</sup> birthday during his paediatric hospitalisation.

The non-MISC cases (n=23) were RT-PCR positive in 22(95.7%); 11(47.8%) were asymptomatic; 12(52.2%) had mild symptoms including fever (30.4%), cough (18.2%) and abdominal pain (21.7%).(Table 2A). Reasons for hospitalization, included bony fracture, haematocolpus, sickle cell disease, chronic kidney disease for haemodialysis, malignancy, appendicitis, diabetic ketoacidosis and four well neonates whose mothers were RT-PCR positive within 48 hours of delivery.

**Table 2A:** Clinical symptoms of children hospitalized with COVID-19, MISC* vs non-MISC.

| Symptoms** | Total cases (%) N= 41 | MISC cases (%) N=18 | Non-MISC cases (%) N=23 | p-Value |
| --- | --- | --- | --- | --- |
| Fever | 24 (58.5%) | 17 (94.4%) | 7 (30.4%) | <0.001 |
| Cough | 8 (20 %) | 4 (22.2%) | 4 (18.2%) | 0.751 |
| Sore throat | 7 (18.4%) | 5 (29.4%) | 2 (9.5%) | 0.116 |
| Rhinorrhoea | 6 (14.6%) | 5 (27.8%) | 1 (4.3%) | 0.035 |
| Shortness of Breath | 7 (17.1%) | 4 (22.2%) | 3 (13%) | 0.438 |
| Chest Pain | 3 (8.1%) | 1 (5.9%) | 2 (10%) | 0.647 |
| Myalgia | 4 (11.1%) | 3 (18.8%) | 1 (5%) | 0.192 |
| Joint Pains | 4 (10.8%) | 3 (17.6%) | 1 (5%) | 0.217 |
| Fatigue/Lethargy | 8 (20%) | 7 (41.2%) | 1 (4.3%) | 0.004 |
| Headache | 4 (10.8%) | 3 (17.6%) | 1 (5%) | 0.217 |
| Seizures | 1 (2.4%) | 0 (0%) | 1 (4.3%) | 0.37 |
| Abdominal Pain/<br>tenderness | 13 (31.7%) | 8 (44.4%) | 5 (21.7%) | 0.121 |
| Vomiting/Nausea | 9 (22%) | 7 (38.9%) | 2 (8.7%) | 0.02 |
| Diarrhoea | 5 (12.2%) | 4 (22.2%) | 1 (4.3%) | 0.083 |
| Conjunctivitis | 6 (14.6%) | 4 (22.2%) | 2 (8.7%) | 0.224 |
| Skin Rash | 9 (22%) | 9 (50%) | 0 (0%) | <0.001 |
| Lymphadenopathy | 6 (14.6%) | 6 (33.3%) | 0 (0%) | 0.003 |
| Bleeding/Bruising | 4 (9.8%) | 3 (16.7%) | 1 (4.3%) | 0.187 |
| Mucositis | 5 (12.2%) | 5 (27.8%) | 0 (0%) | 0.007 |
| Other - signs/symptoms<br>(Anorexia, Arthritis,<br>Irritability, Nasal<br>congestion,<br>Odynophasia, Pedal | 15 (36.6%) | 11 (61.1%) | 4 (17.4%) | 0.004 |
| oedema, Scrotal<br>pain/tenderness |  |  |  |  |

\*Multisystem Inflammatory Syndrome in Children with COVID-19, MISC
\*\*Symptoms with frequency less than one (<4.3), for both groups, were omitted

**Table 2B:**
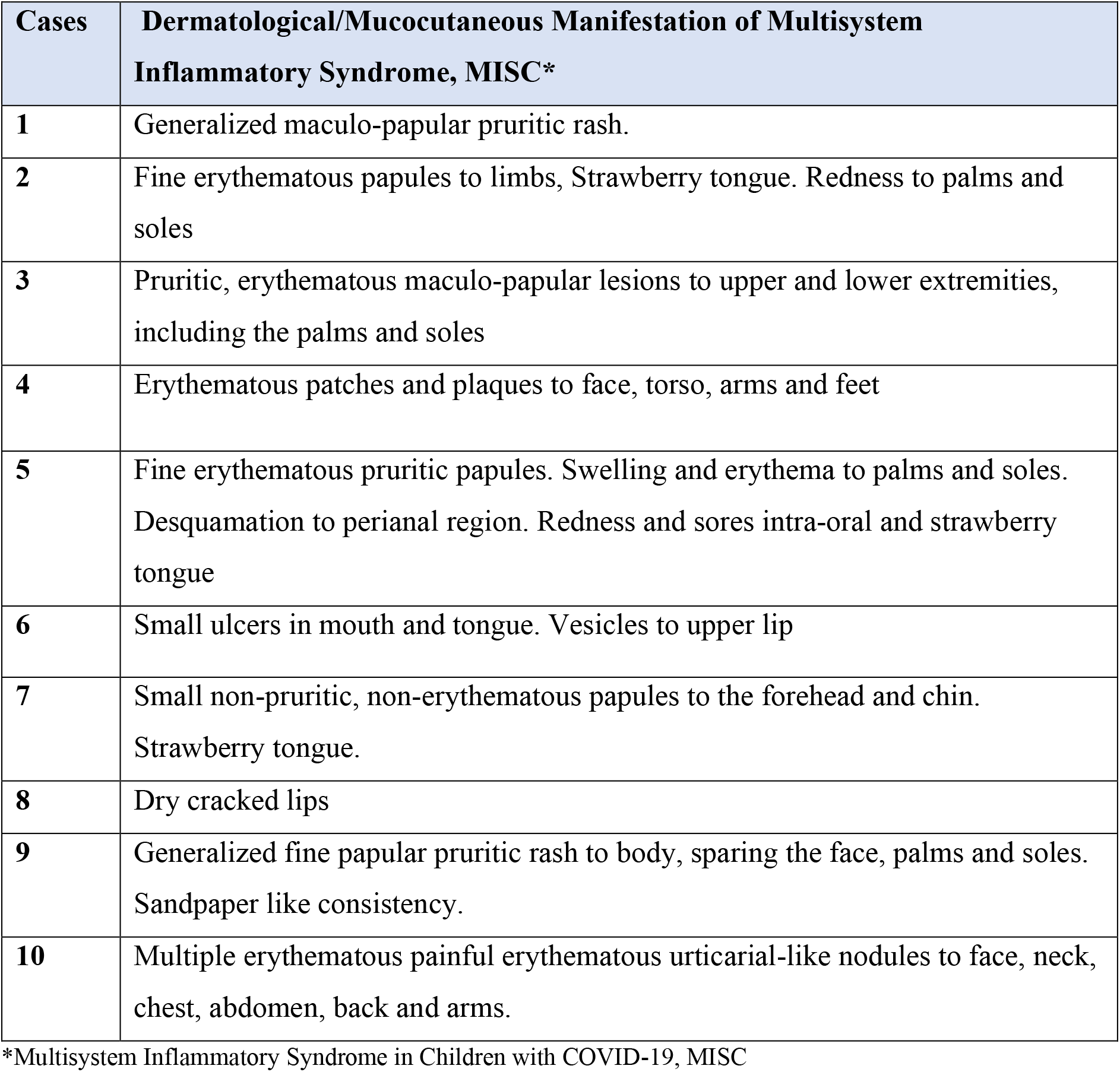
Description of mucocutaneous manifestations in the hospitalized children and adolescents with MISC.

Eighteen (44%) hospitalized cases were diagnosed with MISC: 4(22.2%) having SARS-CoV-2 RT-PCR positive, 7(38.9%) being SARS-CoV-2 IgM/IgG positive and 7(38.9%) having a confirmed/presumed contact. In accordance with CDC’s criteria, 94.4% of MISC patients had fever for ≥24 hours (94% vs. 30% in non-MISC, p<0.001), 100% had ≥2 organ systems involved (18;100% vs. 8; 34.8%, p<0.01)(Table 3); cardiac (17% vs. 0%; p=0.042), with elevated troponin-T (3, 17% MISC), myocardial dysfunction (one, 5.6%), coronary arteritis (one, 5.6%); dermatological (56% vs. 0%;p<0.01) with mucositis (28% vs.

**Table 3:** Organ system involvement in hospitalized children and adolescents with MISC*.

| **Organ system** | **Total cases (%)**<br>**N= 41** | **MISC cases**<br>**(%) N=18** | **Non-MISC**<br>**cases (%)**<br>**N=23** | **p-Value** |
| --- | --- | --- | --- | --- |
| Cases with more than 2 organ involvement | 26 (63.4%) | 18 (100%) | 8 (34.8%) | <0.001 |
| Gastrointestinal | 17 (41.5%) | 13 (72.2%) | 4 (17.4%) | <0.001 |
| Respiratory | 16 (39%) | 9 (50%) | 7 (30.4%) | 0.202 |
| Hematologic | 13 (31.7%) | 12 (66.7%) | 1 (4.3%) | <0.001 |
| Dermatologic | 10 (24.4%) | 10 (55.6%) | (0%) | <0.001 |
| Neurological | 8 (19.5%) | 6 (33.3%) | 2 (8.7%) | 0.048 |
| Other (Rheumatological, Urogenital) | 6 (14.6%) | 5 (27.8%) | 1 (4.3%) | 0.035 |
| Cardiac | 3 (7.3%) | 3 (16.7%) | (0%) | 0.042 |
| Renal | 2 (4.9%) | 2 (11.1%) | (0%) | 0.101 |

\*Multisystem Inflammatory Syndrome in Children with COVID-19, MISC

0%;p=0.007)(Tables 2A,2B); gastrointestinal (13;72% vs. 4;17%; p<0.01) including vomiting/nausea (39% vs. 9%; p=0.02), neurological 33.3%, haematological and coagulopathy (67% vs. 4%;p<0.01), renal 11.1%, respiratory 50% with cough, pleural effusions (17% vs. 0%; p=0.042). Others included urogenital 5.6% and rheumatological 27.8%; Kawasaki Syndrome (44% vs. 0%; p<0.01) and 5.6% (one) demised. MISC cases presented with fatigue/lethargy (41% vs. 4%; p=0.004), lymphadenopathy (33% vs. 0%; p=0.003), rhinorrhoea (28% vs. 4%, p=0.035), other signs/symptoms (61.1% vs 17.4%; p=0.004); including anorexia, odynophagia, nasal congestion, arthritis, scrotal swelling and tenderness and irritability than non-MISC group (Table 2A).

Thirteen (72%) MISC cases had ≥4 abnormal inflammatory biomarkers, including D-dimers, C-reactive protein, ESR, ferritin, troponins, lactate dehydrogenase, neutrophils, platelets, lymphocytes and albumen. The MISC group’s inflammatory markers were more deranged compared to non-MISC group, with ESR (means: 97.07 vs 8.5, p=0.007) and absolute neutrophil count (11.2 v 6.5; p=0.042). Anaemia (17% vs. 0%; p=0.042); neutrophilia (100% vs. 87%; p=0.024) and elevated (≥10mm/hr) ESR (78% vs. 9%; p=0.007) were more likely in MISC cases, with a mean (SD) ESR (mm/hr) 97.07 (38.72) vs 8.5 (6.36) in the non-MISC group (Table 4). MISC cases had more abnormal radiological features, including pulmonary embolism, coronary arteritis, lung consolidation, pleural effusion, pulmonary opacities. Thirteen echocardiograms were done in 72.2% of the MISC group and 11.1% had abnormalities including coronary arteritis, mild mitral regurgitation, elevated pulmonary pressures and a small pericardial effusion in a patient with SCD.(Table 4).

**Table 4:** Investigations of children hospitalized with COVID-19, MISC* v non-MISC.

Investigations of children hospitalized with COVID-19, MISC\* v non-MISC
| Investigations requested** | Total | MISC* | non-MISC | p-Value |
| --- | --- | --- | --- | --- |
| **ESR (Normal <10mmhr)** | 16 (39%) | 14(77.8%) | 2(8.7%) |  |
| Mean | 86 | 97.07 | 8.5 | 0.007 |
| Standard Deviation, SD | 47.089 | 38.72 | 6.36 |  |
| **C Reactive Protein (Normal < 1mg/dL)** | 18 (43.9%) | 16 (88.9%) | 2 (8.7%) |  |
| Mean | 11.4 | 12.2 | 5.1 | 0.512 |
| SD | 13.8 | 14.4 | 7.2 |  |
| **D-dimer (Normal <500 ng/mL)** | 17 (41.5%) | 16 (88.9%) | 1 (4.3%) |  |
| Mean | 6391.8 | 6711.6 | 1275 | 0.652 |
| SD | 11179.5 | 11465.5 |  |  |
| **Ferritin (Normal < 315 ng/mL)** | 13 (31.7%) | 11 (61.1%) | 2 (8.7%) |  |
| Mean | 715.5 | 765 | 443 | 0.634 |
| SD | 829 | 886.8 | 455.4 |  |
| **Lactate dehydrogenase (Normal <190 U/L)** | 20 (48.8%) | 14 (77.8%) | 6 (26.1%) |  |
| Mean | 519.5 | 558.9 | 427.7 | 0.443 |
| SD | 339.1 | 314.3 | 406.9 |  |
| **Neutrophil count (Normal < 8)** | 38 (92.7%) | 18 (100) | 20 (87%) |  |
| Mean | 8.7 | 11.2 | 6.5 | 0.024 |
| SD | 6.5 | 7.9 | 3.9 |  |
| **Lymphocyte count (Normal > 1.5)** | 38 (92.7%) | 18 (100%) | 20 (87%) |  |
| Mean | 2.5 | 1.9 | 3 | 0.056 |
| SD | 1.9 | 1.1 | 2.2 |  |
| **Albumin (Normal: 36 – 52 g/dL)** | 23 (56.1%) | 16 (88.9%) | 7 (30.4%) |  |
| Mean | 34.2 | 33.8 | 35.1 | 0.706 |
| SD | 7.9 | 8.4 | 6.8 |  |
| **Electrocardiogram** | 8 (19.5%) | 8 (44.4%) | 0 (0%) |  |
| -abnormal | 2 (4.9%) | 2 (11.1%) | 0 (0%) |  |
| **Echocardiogram** | 14 (34.1%) | 13 (72.2%) | 1(4.3%) |  |
| -abnormal | 3 (7.3%) | 2 (11.1%) | 1 (4.3%) |  |
| **Abdominal Ultrasound** | 4 (9.8%) | 2 (11.1) | 2(8.7%) |  |
| -abnormal | 3 (7.3%) | 2 (11.1%) | 1(4.3%) |  |
| **Computerised Tomogram of the Chest** | 2 (4.9%) | 2 (11.1%) | 0 (0%) |  |
| -abnormal | 2 (4.9%) | 2 (11.1%) | 0 (0%) |  |
| **Computerised Tomogram of the Abdomen** | 3 (7.3%) | 1 (5.6%) | 2 (8.7%) |  |
| -abnormal | 1 (2.4%) | 0 (0%) | 1 (4.3%) |  |
| **Lower Limb Doppler Ultrasound** | 2 (4.9) | 2 (11.1%) | 0 (0%) |  |
| -abnormal | 0 (0%) | 0 (0%) | 0 (0%) |  |
| **Chest Radiograph** | 10 (24.4%) | 6 (33.3%) | 4 (17.4%) |  |
| -abnormal | 6 (14.6%) | 5 (27.8%) | 1(4.3%) |  |

\*Multisystem Inflammatory Syndrome in Children with COVID-19, MISC
\*\* Fibrinogen, Interleukin 6, Procalcitonin were not available

Patients from both groups were treated for their complaints (Table 5). Fourteen of the MISC group received intravenous gammaglobulin (IVIgG) 14(77.8%) vs 0; p< 0.001), 12(66.7%) received aspirin and 9(50%) high dose corticosteroids. Others included appendectomy 2(8.7%) in non-MISC cases and antibiotics 15(83.3%) MISC vs 13(56.5%) non-MISC; p=0.067). Mean (SD) duration of admission for MISC and non-MISC groups were 12.4 (8.8) and 10.55(18.6%); p=0.693, respectively.

**Table 5:** Medical Intervention and outcomes of children hospitalized with COVID-19, MISC^*^ v non-MISC.

| **Interventions and Outcomes** | **Total cases (%)**<br>**N= 41** | **MISC\* cases**<br>**(%) N=18** | **Non-MISC**<br>**cases (%)**<br>**N=23** | **p-Value** |
| --- | --- | --- | --- | --- |
| Intravenous Immunoglobulins (IVIgG) | 14 (34.1%) | 14 (77.8%) | 0 (0%) | <0.001 |
| Anticoagulants | 7 (17.1%) | 5 (27.8%) | 2 (8.7%) | 0.107 |
| Aspirin, ASA | 12 (29.3%) | 12 (66.7%) | 0 (0%) | <0.001 |
| Antiviral | 2 (4.9%) | 2 (11.1%) | 0 (0%) | 0.101 |
| Broad Spectrum Antibiotics | 28 (68.3%) | 15 (83.3%) | 13 (56.5%) | 0.067 |
| Corticosteroids | 11 (26.8%) | 9 (50%) | 2 (8.7%) | 0.003 |
| Complications | 12 (29.3%) | 11 (61.1%) | 18 (78.3%) | 0.231 |
| -Anemia | 3 (7.3%) | 3 (16.7%) | 0 (0%) | 0.042 |
| -Pleural Effusion | 3 (7.3%) | 3 (16.7%) | 0 (0%) | 0.042 |
| -Pulmonary Embolism | 1 (2.4%) | 1 (5.6%) | 0 (0%) | 0.252 |
| -Cardiac Arrest | 1 (2.4%) | 1 (5.6%) | 0 (0%) | 0.252 |
| -Cardiac Arrhythmia | 0 (0%) | 0 (0%) | 0 (0%) | - |
| -Cardiomyopathy | 0 (0%) | 0 (0%) | 0 (0%) | - |
| -Coronary arteritis | 1 (2.4%) | 1 (5.6%) | 0 (0%) | 0.252 |
| -Pneumonia | 5 (12.2%) | 3 (16.7%) | 2 (8.7%) | 0.439 |
| -Acute Respiratory Distress Syndrome, ARDS | 2 (4.9%) | 2 (11.1%) | 0 (0%) | 0.101 |
| Duration of Hospitalization, days; Mean (SD) | 11.41 (14.9) | 12.4 (8.8) | 10.55 (18.6) | 0.693 |
| Admission to Intensive Care Unit | 2 (4.9%) | 2 (11.1%) | 0 (0%) | 0.101 |
| Transferred | 1 (2.4%) | 1 (5.6%) | 0 (0%) | 0.252 |
| Discharged | 39 (95.1%) | 16 (88.9%) | 23 (100%) | 0.101 |
| Demised | 1 (2.4%) | 1 (5.6%) | 0 (0%) | 0.252 |
| Readmitted | 2 (4.9%) | 2 (11.1%) | 0 (0%) | 0.101 |
| Readmitted --- received Supportive Care | 2 (4.9%) | 2 (11.1%) | 0 (0%) | 0.101 |
| Readmitted --- received second IVIgG and IV Steroids | 1 (2.4%) | 1 (5.6%) | 0 (0%) | 0.252 |
| Readmitted --- Discharge | 2 (4.9%) | 2 (11.1%) | 0 (0%) | 0.101 |
| Readmitted --- Demise | 0 (0%) | 0 (0%) | 0 (0%) |  |

\*Multisystem Inflammatory Syndrome in Children with COVID-19, MISC

Notable cases include one who presented during the second month of SARS-CoV-2 in Jamaica with dyspnoea, lethargy, hypercoagulability, systemic inflammation, negative bacterial cultures and anaemia (nadir 2.8 g/dL) requiring packed red blood cell transfusions. Her computerised chest tomogram showed “ground glass” opacities in both lungs; “crazy paving” and “consolidation bands” involving 50% to 75% of right lung and 25% of the left lung. She had deep vein thrombosis involving the lower limbs. She received anticoagulation, steroids, antibiotics, antivirals and non-invasive ventilation. SARS-CoV-2 RT-PCR and serology were negative and she had close interaction with a confirmed positive contact. Another child, hospitalized one month after Jamaica declared community transmission for SARS-CoV-2, presented with prolonged fever, persistent diarrhoea and irritability, elevated erythrocyte sedimentation rate [ESR], d-dimers, thrombocytosis, monocytosis. His entire household was SARS-CoV-2 RT-PCR positive, two weeks prior to his symptoms, his RT-PCR (x3) and serology (Abbott, nucelocapsid IgG test) were negative. His admission sample was retested 6 weeks later and the SARS-CoV-2 spike protein IgG was positive. Two children with Sickle Cell Disease (HbSS) were; one had fever, rash, abdominal pain, respiratory symptoms, elevated/or abnormal inflammatory markers and SARS-CoV-2 RT-PCR positivity 4 weeks prior. She was treated with IVIgG, intravenous steroids, aspirin and improved. However, two weeks later she was readmitted with headaches, unilateral weakness of the lower limb and worsening systemic inflammation requiring exchange transfusion, a second dose of IVIgG, intravenous corticosteroids and long-term anticoagulation. Her weakness resolved by day 3 of the second admission. The other patient with HbSS disease had bilateral pulmonary consolidation, right pulmonary artery thromboses, right lung abscess and empyema, elevated inflammatory markers and he was SARS-CoV-2 IgG positive. He also responded to IVIgG, high dose IV steroids, aspirin, anticoagulation, broad-spectrum antibiotics, thoracostomy and ambulatory anticoagulation therapy. Another child, with leukaemia, was readmitted ten days after initial hospital discharge for MISC, with altered mental status and delirium, resolving by day three of the readmission. A neonate who was born to a SARS-CoV-2 positive mother had prolonged respiratory distress, was small for gestational age and was hospitalized. He has significant neuro-developmental challenges and autoimmune myasthenia gravis.

Thirty-nine (95.1%) of 41 hospitalised children were discharged to follow-up. One (2.4%) patient with MISC commenced IVIgG, corticosteroids and aspirin and was transferred to home country and completed treatment. There was one death (2.4%) from sudden myocardial dysfunction 13 days after testing RT-PCR positive, in a child with HbSC disease receiving haemodialysis for pre-existing chronic kidney and bony metabolic disease.

## DISCUSSION

Forty one (52%) children and adolescents with SARS-CoV-2 infection and/or MISC were hospitalized consecutively, from 79 children who presented to UHWI with SARS-CoV-2 infection, during April 2020 through June 2021. Careful surveillance recognized 44% had MISC when the CDC’s criteria were consistently and conscientiously applied. This shows that MISC maybe common. Children from all age groups studied, presented with SARS-CoV-2 infection, however those over six years of age were more likely to have MISC. These occurred while COVID-19 and MISC were being originated and globally defined and as treatment recommendations evolved. Hence, this case series captures children hospitalized during the COVID-19 pandemic in its first fifteen months of evolution in one regional academic medical center in a developing country from the Caribbean.

Most patients did well. They did not have fluid refractory shock, or require inotropes, vasopressors, volume expanders, or invasive ventilation. Only 11.1% (2) MISC vs 0% non-MISC cases required intensive care, for non-invasive ventilation. This is significantly less than in other reports. In a Southern Turkey MISC series, 17.3% of children needed inotropes and 28.8% intensive care (Toulunay et al., 2020). In a New York multi-institutional study, 51% received vasopressors and 15% mechanical ventilation (Kaushik, 2020). In a large cohort study in the USA, 54% had hypotension, or shock and 58.2% required intensive care (Belay et al., 2021). In Spain, a multi-center study reported 84.4% MISC vs 13.8% non-MISC children in shock and 13.3% MISC vs 41.4% non-MISC required invasive ventilation (Garcia-Salido et al., 2020). From England, 50% were in shock while 79% required invasive ventilation (Whittaker et al., 2020). While a case series in Qatar had 71.4% requiring inotropes in intensive care (Hasan et al., 2020). In the developing world, reports from Mexico and Pakistan, had 23% and 25% children with MISC in shock, respectively (Macias-Parra et al., 2021; Sadiq et al., 2020). A meta analysis of MISC cases revealed 65.8% developed shock, 79.1% required admission to the intensive care unit and 33.0% required mechanical ventilation (Yasuhara J et al, 2021).

The improved outcomes in our study may be multi-factorial. There was a high index of suspicion by clinicians, from widespread educational sessions, on COVID-19 and MISC, early and throughout the pandemic; knowledge and use of the CDC’s MISC diagnostic criteria which are simple to implement. Paediatric infectious disease consultation in our institution, led to early referrals to and in our hospital, prompt diagnosis and aggressive treatment, with IVIgG, aspirin, intravenous steroids and monitoring of inflammatory markers which likely led to more favourable outcomes (discharged MISC vs. non-MISC 94.4% vs 100%). In our study, IVIgG was given to 77.8% of our MISC cases and 50% of our MISC patients also received IV glucocorticoids. This may be beneficial in resource-constrained settings, such as Jamaica. However, IVIgG is costly and may affect accessibility and outcomes in children as the pandemic surges. A study from the United States, reported initial treatment with IVIgG and intravenous glucocorticoids at diagnosis of MISC, led to reduced left ventricular dysfunction and need for vasopressors than if IVIgG was given alone (Son et al, 2021). Measures must be implemented to ensure that all children have equal access to these life-saving medications once MISC is diagnosed.

The ethnic diversity of our population may also be protective. A prospective cohort study in the US reported Black and Hispanic race as predictors for hospitalization in children with COVID-19, with 19.5% of hospitalized children required intensive care (Howard LM et al., 2021). Apolipoprotein E4 (apoE4) may be present in high frequency in individuals of African descent compared to Europeans or Asians and this may be a predictor of rapid and severe illness in COVID-19, as it leads to robust immune responses (Goldstein et al., 2020; Goldstein et al., 2020). This may contribute to the differences in severity between the races. Jamaica is ethnically diverse because of the many populations that initially settled in the region and the large, growing diaspora (Premdas et al., 2011). The ethnicities of Caribbean countries include Europeans, Africans, Asian Indians, Indonesian Javanese, Chinese, Aboriginal Indians and many mixes (Premdas et al., 2011). No other region of the world may be so richly diverse (Premdas et al., 2011). However, there are no reports characterizing the ethnic descent of the populace of each island. This ethnic diversity may have an added protective effect, against the variants during the study.

The outcomes in our study may be linked primarily to the B.1.1.7 (alpha, UK) SARS-CoV-2 “variant of concern” and other minor variants, which were circulating in Jamaica during the period of our study (Charles J, 2021; COVID-19 Ministry of Health, Jamaica, 2021). This may be equated to the experiences in other countries, although most reported more severe outcomes during the same 15-month time period of the global pandemic. More severe clinical manifestations and outcomes may emerge in Jamaica, with circulation of the B1.617.2 (Delta, Indian) and other SARS-CoV-2 “variants of concern” (PAHO/WHO, 2021).

Children with MISC had a wide array of organ-system involvement and abnormal inflammatory markers. Both simple, more accessible inflammatory markers, like complete blood count, lactate dehydrogenase, ESR and CRP, as well as D-dimer, ferritin and troponins, which are more costly and less accessible in the developing world setting, were available to our patients and were abnormal. This supports the multi-system, inflammatory nature of MISC (Ahmed et al., 2020; Aranoff et al., 2020; Consiglio et al., 2020; Feldstein et al., 2021; Hoang et al., 2020; Howard et al., 2021; Radia et al., 2020). Just under a quarter of the MISC cases were RT-PCR positive and of the RT-PCR negative MISC cases, 33.3% had positive IgG/IgM antibodies and the remaining 38.9% had confirmed, or presumed contact, especially during widespread and intense community transmission. This is congruent with the post infectious nature of MISC. Many institutions in Jamaica do not have access to SARS-CoV-2 antibody testing, despite its availability to our patients in this UHWI study. The lack of testing could lead to evidence of SARS-CoV-2 being missed and families may be unaware of their child’s exposure or acute infection, which maybe asymptomatic. Despite these challenges, the CDC’s diagnostic criteria in a period of intense community transmission, since September 2020 through June 2021, should assist clinicians in making the diagnosis and implementing treatment since community transmission increases the likelihood of SARS-CoV-2 exposure. Antibody testing, as well as a high index of clinical suspicion, may reduce delays in diagnosis and treatment of MISC. A US study, reported that MISC followed their COVID-19 community peaks by two to five weeks (Belay et al., 2021). All patients in our study identified clinically as MISC without serological evidence (38.9%), had multisystem involvement and abnormal inflammatory markers, without another aetiology to explain their symptoms. All the children treated for MISC had improvement of their clinical findings and inflammatory markers once therapy was started.

Cardiac decompensation, though present in under one fifth of our sample population was not a significant feature in our study, in contradistinction to other reports (Ahmed et al., 2020; Aranoff et al., 2020; Consiglio et al., 2020; Feldstein et al., 2021; Hoang et al., 2020; Howard et al., 2021; Radia et al., 2020, Yasuhara J et al, 2021). Our findings may be attributed to the prompt diagnosis and treatment, variant strain, or the ethnicity (Hoang et al., 2020; Howard et al., 2021; Son et al., 2021). Challenges in accessing echocardiogram and/or electrocardiogram in the acute setting may have also led to radiological under-representation. However, our patients did not have clinical cardiac decompensation and troponins, when performed, were not frequently abnormal. One patient, with multiple co-morbidities including HbSC disease and chronic renal failure requiring haemodialysis, had sudden cardiac dysfunction and demised 13 days after his positive RT-PCR result. Globally, co-morbidities have been noted to lead to more severe outcomes (Belay et al., 2021; Garcia-Salido et al., 2020; Howard et al., 2021; Kaushik et al., 2020; Macias-Parra et al., 2021; Sadiq et al., 2020; Son et al., 2021; Tolunay et al., 2020; Whittaker et al., 2020). Children should therefore be prioritised for COVID-19 vaccinations, once authorized, accessible and available for their age groups.

## Limitations

Some investigations were not readily available in the acute setting. However, 72.2% of patients with MISC obtained echocardiograms done later and very few had cardiac involvement. Some patients did not return, or respond to telemedicine follow-up visits.

## Conclusion

In this study, more than half of the children aged <16 years with SARS-CoV-2 infection presenting to the UHWI during the first 15 months of the epidemic were hospitalized. Children without MISC had a mild course. Forty-four percent (44%) of the hospitalized were promptly diagnosed with MISC and 94% of these were successfully treated with IV gammaglobulin, steroids and/or aspirin before cardiopulmonary decompensation and need for inotropes, vasopressors, or invasive ventilation. One death from myocardial dysfunction occurred in a child with pre-morbid Sickle Cell Disease and chronic renal failure on haemodialysis. Children and adolescents with co-morbidities should be prioritised for SARS-CoV-2 vaccinations. A high index of suspicion, knowledge and correct use of the MISC diagnostic criteria, with prompt treatment may lead to improved outcomes in children in resource scarce countries.

## Data Availability

All data produced in the present work are contained in the manuscript

## AUTHOR RESPONSIBILITIES

(ICMJE criteria, updated December 2019):

1. “Make contributions to the conception or design of the work; or the acquisition, analysis, or interpretation of data for the work” (CSB and CDCC concepted and designed the work; CSB, RHC, ANH, JJA, CDCC acquired the data; CLB, KMG and VMD performed data analysis and everyone commented on the proposal, the work and data interpretation throughout); AND
2. “Drafting the work, or revising it critically for important intellectual content” (CSB and CDCC drafted and revised the work, all others contributed to the process throughout); AND
3. “Give final approval of the version to be published” (everyone approved the final version submitted for publication); AND
4. “Be accountable for all aspects of the work in ensuring that questions related to the accuracy or integrity of any part of the work are appropriately investigated and resolved” (all co-authors agreed to be responsible for all aspects of the work reported herein).

## CONFERENCE PRESENTATION

An abstract was published and a poster presentation of this manuscript has been displayed and was virtually presented to the American Academy of Paediatrics (AAP), National Conference and Exhibition, on October 8 -12, 2021 in the Section on “Global Health Program” (VH1309), at the Philadelphia Convention Center, Pennsylvania, USA.

## ACKNOWLEDGEMENTS

The authors acknowledge the support Dr David McGrowder and the Chemical Pathology Laboratory of the University Hospital of the West Indies for performing highly specialised investigations, including inflammatory markers. Dr Reayah Francis is acknowledged for assistance with data entry and Mrs Charmaine Neath-Hewitt for administrative assistance. We are grateful and appreciative to the entire health care team of the University Hospital of the West Indies, especially the paediatric consultants, residents, nurses and pharmacists who assisted in providing holistic care of these children. Finally, we thank the affected children and families and hope our interventions enabled them to improve their quality of life.

